# Examining clan conflict dynamics and their role in driving displacement: insights from Somalia

**DOI:** 10.1101/2025.10.30.25339180

**Authors:** Mohamud Isse Yusuf, Ahmed Adan Mohamed, Abdisamad Abdirahman Omar

## Abstract

This study examined the clan conflict dynamics and their role in driving displacement in Somalia. A comprehensive literature review and quantitative retrospective data analysis were employed to obtain the study’s data. The study used inferential statistical tests of correlation and linear regression analysis to examine the association between clan conflict frequency and displacement. Utilizing correlational analyses, the research demonstrates a strong positive correlation (Pearson’s r = 0.705, p < 0.05) between clan-based conflict and displacement.

Moreover, the study examines the relationship between clan-based conflicts and population displacement in Somalia using Linear Regression Analysis. The results indicate a robust positive association between these two variables, with a correlation coefficient (R) of 0.705. The statistical significance of the regression model is confirmed by the ANOVA results, which show an F-statistic of 8.891 and a p-value of 0.015, demonstrating that clan conflict serves as a significant predictor of displacement. The coefficients table further corroborates this finding, revealing a standardized coefficient (Beta) of 0.705 and a p-value of 0.015, which suggests a strong and statistically significant association between clan conflict and displacement. Finally, the study proposes several recommendations to address these issues, including community-level resilience initiatives, early conflict warning systems, data-driven decision-making, sustainable income generation programs, and multi-stakeholder collaboration.

## Introduction

Globally, conflict is seen as a key issue that impedes growth (Happi, 2016). Throughout history, numerous regions worldwide, particularly arid areas with limited natural resources, have been plagued by conflicts. Huho (2012) suggests that conflict arises from structural inequalities and imbalanced power distribution. A conflict situation emerges when two or more distinct groups actively confront each other, pursuing incompatible objectives (Salad, 2015). Other scholars also suggest that conflict emerges from competing interests over limited resources, divergent goals, and frustration. This occurs in relationships characterized by both competitive and cooperative elements, where individuals have conflicting motivations (Kamta & Scheffran, 2022). However, Conflict is an unavoidable aspect of society, emerging through various processes such as societal shifts, individual psychological growth, economic disparities, the formation of cultural identities, and the structuring of political systems (ONGAGA, 2017).

Conversely, global displacement patterns have been increasing due to various challenges including climate change, insecurity, and conflict. This worldwide surge in population displacement has significantly affected the global food system (Osman & Abebe, 2023). According to the United Nations, roughly 30% of the world’s population, primarily in developing nations, is currently facing food insecurity. Conflicts leading to mass migration are a major contributor to these issues. The forced abandonment of agricultural lands disrupts food production and frequently results in famine (Osman & Abebe, 2023).

For the past 25 years, Somalia has been devoid of any reliable peace and stability since 1991 (Mohamed & Samantar). It is widely accepted that the ongoing conflicts driven by political, clan, and religious factors in Somalia have resulted in a significant wave of displacement, both within the country and as refugee migration to other nations(Mohamed & Samantar). Most conflict-related research often neglects the connections between development, environmental, and conflict-driven displacements.

However, in Somalia’s case, four interconnected factors have contributed to population displacement: armed conflict, ethnic and racial persecution, environmental disasters like drought, floods, and famine, and development projects altering land use and ownership (Fellin, 2013). In this instance, a significant portion of Somalia’s population has been displaced, both within and outside the country, due to violence, conflict, famine, severe food shortages, and the resulting poverty and instability (Robinson et al., 2014). For instance, Dadaab in Northeastern Kenya hosts the world’s largest refugee camp, containing the highest concentration of Somalis outside of Mogadishu. Somalia has been characterized not only as experiencing one of the world’s most severe human rights and humanitarian crises but also as having one of the most critical displacement situations globally(Robinson et al., 2014). From an operational perspective, Somali refers to ethnic pastoral communities residing in Somalia, Kenya, and Ethiopia, who share similar cultural practices, linguistic heritage, and religious beliefs (Alio, 2012).

### LITERATURE REVIEW

Conflict, in its simplest form, occurs when individuals or groups struggle over limited supplies of tangible or intangible goods (Shale, 2011). Moreover, conflicts among communities are primarily driven by the struggle for control and access to natural resources, especially water and grazing lands. Additional factors contributing to these conflicts include long-standing rivalries, deeply ingrained cultural beliefs, land disputes, political instigation, youth unemployment, and the recent increase in illegal weapons (Salad, 2015).

Some researchers point out that the exploitation of natural resources and related environmental pressures play a significant role in all stages of conflicts, from their inception and continuation to hindering peace efforts. They have observed that natural resources are linked to at least 40% of internal conflicts over the past six decades. The repercussions of conflict on communities are extensive, causing severe and enduring damage to their social, economic, and political landscapes. When civilians become direct targets or unintended victims, the social fabric and coping mechanisms of a society are torn apart, making the return to normalcy a prolonged process that can span years. This is particularly true when social institutions and ways of life are intentionally destroyed(Salad, 2015). During times of strife, extended family and kinship networks offer crucial support. However, conflict also intensifies divisions between groups, escalates intra-group tensions and mistrust, disrupts economic relationships between different factions, and creates conditions conducive to disease spread. A stark example of this can be seen in Rwanda, where even after the violence subsided in mid-1994, deaths continued as refugees and internally displaced persons fell victim to diseases caused by insufficient food and clean water. As a result, the political, economic, and social frameworks of conflict-affected communities undergo significant transformations (Salad, 2015).

The Horn of Africa is characterized by persistent violent conflicts occurring at multiple levels: national, regional, and local. These conflicts involve a diverse range of participants, including governments, nationalist organizations, religious factions, and community or identity groups, often with substantial support from external sources(A. A. Mohamed, 2018). A notable feature of these conflicts is their tendency to quickly expand beyond national boundaries, taking on a sub-regional dimension. This cross-border nature is facilitated by the involvement of ethnic kin from neighboring countries within the Horn of Africa region (A. A. Mohamed, 2018).

For nearly 30 years, Somalia has experienced a lack of central government due to a combination of factors, including civil war, political instability, and clan-based armed conflicts. These issues stemmed from efforts to overthrow the dictatorial regime and subsequently led to a power vacuum and unrest, particularly in the southern region. Somalia’s society is traditionally organized around clans, which have their methods for addressing inter-clan matters such as disputes, power distribution, and criminal offenses committed by clan members. The conflict in Somalia stems from a variety of intricate factors, encompassing political, economic, cultural, and psychological aspects. Throughout the different phases of the conflict, numerous internal and external parties have exerted varying degrees of influence (Ahmedweli Mohamud, 2012). These methods include the use of law enforcement and judicial systems, intermarriage, and other conflict-resolution mechanisms (Barrow, 2020).

Historically and since the 1800s, Somalia has experienced inter-clan disputes stemming from competition over essential resources like water, livestock, and pastures. Historically, Somali nomads have engaged in conflicts over camel ownership due to the animals’ crucial role in surviving Somalia’s challenging environment (Abdirahman, 2016). In this setting, clan affiliation proved valuable, as amassing and maintaining a substantial camel herd required the backing of one’s clan members. During the initial phase of the civil war (1988-1992), militias were organized along major clan divisions, resulting in the frequent exchange of control over major urban centers. At that time, it was common for media outlets and Somali people to report that a specific clan had seized control of a particular city (Abdirahman, 2016). A civil war occurs when organized factions within a single nation-state or republic engage in armed conflict, or less frequently when two countries that were previously part of a unified nation-state fight each other (Abdikarim Mohaidin, 2011). Furthermore, Somalia has emerged as the most notable case of state collapse in Sub-Saharan Africa over recent years (Jama et al., 2018).

In southern Somalia, violent clashes erupted among clan-based armed groups vying for dominance over important urban centers, ports, and residential areas. The primary casualties of this conflict were vulnerable farming communities and minority groups residing along the coast, who found themselves caught in the crossfire of the warring factions (Abdikarim Mohaidin, 2011). The Somali conflict encompasses all major Somali clans, including Hawiye, Darod, Isaq, Dir, Rahanweyn, and nearly every minority group. Each clan either operates its faction or participates in a coalition of factions. Certain groups have exerted dominance over others, compelling the subjugated to seek alternative means of survival in Somalia’s lawless environment. This dynamic has resulted in every clan becoming an active participant in the ongoing conflict (Farah et al., 2002).

Understanding the political structure of Somali society hinges on grasping the concept of kinship and its unique social contract. As long as Somalis rely on their kinship lineage for protection and security, collective rather than individual perspectives will continue to shape responsibilities, obligations, rights, and liabilities. Consequently, clans will remain collectively accountable for the actions of their individual members, while the rights of women and children will persistently be viewed through the lens of preserving the strength of male-dominated clans (Gundel, 2009).

The Somali clan system is founded on a patrilineal kinship structure. Clan membership is determined by tracing one’s ancestry through male lineage. The social agreement outlines the principles of unity within and among these patrilineal clans. An analytical breakdown of the traditional Somali structures reveals three fundamental components: 1) The segmentary lineage system, which forms their traditional social organization; 2) The xeer, which comprises their customary laws; 3) Their traditional juridic-political framework, represented by traditional authorities. Genealogical records establish an individual’s clan affiliation based on their ancestral origins (Gundel, 2009).

Worldwide, internal displacement stands as the predominant form of forced migration and serves as a crucial tactic in eliminationism politics. Migration experts typically employ a framework centered on rational choice and utility maximization to examine patterns of migrant movement. Seminal models depict individuals assessing the expenses of departing against the potential benefits of relocating to various destination nations (Blair, 2024). This decision-making process, which determines whether, when, and where to migrate, is subject to uncertainties and financial constraints. The factors prompting individuals to leave their homes are categorized as “push” factors, while those drawing them toward specific destinations are termed “pull” factors. Despite extensive testing of this migrant decision-making model in the context of international migration, there remains a dearth of systematic analysis regarding internal displacement patterns(Blair, 2024).

With the seventh-largest internally displaced population globally, Somalia has been the focus of one of the world’s most enduring humanitarian aid efforts since the late 1980s (Drumtra, 2014) and in the meanwhile, prior to the collapse of the previous central government in 1991, Somalia was already ranked among the world’s most impoverished nations, with an unstable economy that relied heavily on livestock trade(Mohamed & Samantar). In 2023, a record 2.9 million people were forced from their homes due to environmental disasters and conflict. Climate-related events accounted for 75 percent of displacements, affecting 2.3 million individuals (1.7 million from floods and 531,000 from drought). Concurrently, conflict and insecurity led to the displacement or re-displacement of 653,000 people, marking an unprecedented high. Operational challenges impact over 3.8 million people, with approximately 580,000 residing in areas that humanitarian organizations struggle to reach. Most of those in hard-to-access regions are women and children(UNOCHA, 2024). Conversely, the clan conflicts and forced displacement have resulted in significant loss of life, disrupted the livelihoods of millions of internally displaced persons (IDPs) and their host communities, and caused extensive damage to infrastructure. Furthermore, many IDPs report that the food aid provided by humanitarian organizations is inadequate to meet their needs (Sate & Konso Zone).

Furthermore, as of late 2022, approximately four million individuals remained internally displaced within the nation. Both armed conflicts and climate-related issues have prompted population movements, including exodus to refugee camps in adjacent countries, repatriation to Somalia from foreign lands, or internal displacement (Addow, 2024). This displacement often stems from extended periods of hostilities, strife, and climate-induced events that undermine livelihoods, exhausting all available resources and adaptive strategies. Nevertheless, mobility can also represent a more deliberate choice, shaped by multiple factors, as people pursue enhanced prospects for themselves and their families(Addow, 2024), conflict also hampers agricultural activities, exacerbates food shortages, and triggers population displacement, including the formation of camps for internally displaced persons (IDPs) (Kinyoki et al., 2017).

The origins of displacement in modern-day Somalia can be traced to former colonial divisions, clan rivalries, and intense competition for economic and political power in the post-colonial era. Somalia’s independence in 1960 was hindered by a lack of political credibility and a fragile economic foundation (Mustafe Mahamoud, 2017). The government’s seizure of economic assets, the impact of a severe drought in 1974, and the military defeat in the 1978 Ogaden conflict led to widespread disillusionment with the Barre administration. Serious armed resistance against Barre began in 1988 in northwestern Somalia, causing approximately 400,000 Somalis to seek refuge in Ethiopia and Djibouti. Barre’s ousting in 1991 plunged Somalia into an extended period of civil strife. During the peak of the conflict in 1992, around 800,000 Somalis were refugees in adjacent countries, while 2 million were displaced within Somalia’s borders(Mustafe Mahamoud, 2017).

The displacement has become one of the most significant crises facing the world in modern times(Yigzaw & Abitew, 2019). Displacement resulting from conflict can significantly alter an individual’s connection to their community. This forced relocation extends beyond merely increasing the physical distance between a person and their familiar surroundings (Aymerich & Zeyneloglu, 2023). It compromises the displaced individual’s capacity to contribute to community decisions, fully participate in communal affairs, interact through established norms and values, and engage in shared experiences that collectively define one’s Sense of Community (SOC). Moreover, the economic hardships associated with displacement, combined with the destruction of infrastructure due to conflict, further reduce the material resources individuals can dedicate to and invest in their community (Aymerich & Zeyneloglu, 2023).

The sense of community (SOC) experienced by members toward their original community may be diminished due to damage and separation. However, the shared ordeal of conflict and forced relocation could serve as a powerful bonding experience that strengthens SOC. Internally displaced persons (IDP) often experience nostalgic feelings for their original community, stemming from the identity disruption caused by forced displacement (Aymerich & Zeyneloglu, 2023). Struggling to adapt to unfamiliar surroundings, IDPs tend to reinforce their collective community norms and traditions. The yearning for familiar community practices may act as a driving force, leading to increased levels of SOC. This dual dynamic, where displacement both supports and undermines community members’ attachment to their original community, raises significant questions about the SOC levels experienced by forcibly displaced individuals in unstable environments (Aymerich & Zeyneloglu, 2023).

Examination of data on daily population movement and incidents of violence reveals that structural elements, particularly shifts in the geographic range and power dynamics of conflicts, are the primary factors influencing increases or decreases in displacement. These impacts are a result of the proportion of people affected by the conflict, disruptions to security, and widespread fear. The study makes two key contributions (Schon, 2015). Firstly, it highlights the intricate nature of the relationship between violence and displacement. Secondly, it underscores the significance of considering the structural aspects of conflicts, even as advancements in data collection and methodology have enabled researchers to focus on more granular details of these conflicts(Schon, 2015).

Examining the impact of clan-driven displacement on livelihoods, there are worries about how long-term conflict and forced relocation diminish household resources. This has constrained the ability of impoverished Somalis to support vulnerable children, as entire families often share the food rations intended for child patients (Le Sage & Majid, 2002). Other contributing factors include inadequate water and sanitation facilities, as well as poor nutritional quality even when food is available. Moreover, some households experience extremely slow recovery, with many becoming destitute. Ongoing political and economic exclusion may prevent certain social groups, including minorities and small sub-clans, from accessing markets (Le Sage & Majid, 2002).

According to Collins (2004), the term clan is conceptualized as “an informal organization consisting of individuals related or linked by kin-based bonds.” The core of a clan is comprised of emotional kinship connections, which form the foundation of its organizational identity and relationships. These connections extend both vertically and horizontally, connecting leaders with followers, and encompass both genuine blood relations and symbolic kinship. The etymology of the word clan can be traced back to “clan” in Irish and Scottish Gaelic languages, which translates to family (Hussein, 2022). In contrast, conflict is conceptualized here as the dynamic between interdependent individuals who perceive their aims as clashing and view each other as obstacles in the pursuit of these objectives(Jibril, 2013). It’s also worth mentioning that clan conflict is a type of conflict that occurs within a clan (A. Mohamed, 2018) while a person who is compelled to leave their residence but remains within the boundaries of their nation is known as an Internally Displaced Person (IDP)(Muna Mohamed, 2020).

The study is grounded in Karl Marx’s conflict theory, which posits that society exists in a constant state of discord due to the struggle for scarce resources. This theory suggests that social order is maintained through dominance and power, rather than agreement and compliance (Adan & Orodho, 2013). Conflict theory argues that individuals in positions of wealth and influence strive to maintain their status by any means necessary, primarily through the oppression of those without power. Furthermore, the theory contends that major historical advancements, such as the establishment of democratic systems and civil liberties, stem from capitalist efforts to control the populace, rather than from a genuine desire for societal harmony (Adan & Orodho, 2013).

This conflict theory has been applied to elucidate various social issues, such as warfare, uprisings, economic disparities, resolve, and family abuse. Societies are composed of groups that necessitate rules. Groups can be chaotic entities. In the absence of guidelines, individuals tend to deviate, either unintentionally or purposefully. A social mechanism is essential to function as a herding dog, steering people back to the collective (Adan & Orodho, 2013).

The research also referenced Henri Tajfel’s social identity theory from the 1970s, which uses the concept of social identity to explain prejudice, discrimination, and conflict between groups. According to this theory, individuals first develop preferences for certain groups, then align themselves with these favored groups (known as “in groups”), and finally compare themselves to other groups perceived as “out groups.” This process is essential as social groups aim to differentiate themselves from one another. Members of clans typically prefer to identify with their birth clan and resist being recognized as part of other clans, even when living in the same village, due to concerns about losing their clan identity. People often describe the external category using falsehoods, exaggerated labels, and negative attributes, while associating their own group with positive characteristics (Sahal, 2016).

Thus, despite extensive research on conflict dynamics and displacement in Somalia, significant knowledge gaps persist regarding the complex interrelationship between clan-based conflicts and displacement trends. Current scholarly literature predominantly focuses on the historical and structural elements of clan rivalries, competition for resources, and the sociopolitical ramifications of displacement. However, a comprehensive analysis of how these elements interact to focus on relationship of these variables (clan conflict and displacement) is lacking. Furthermore, while some studies emphasize the role of scarce natural resources and political instigation in precipitating conflicts, these investigations fail to provide a thorough examination of the correlation between clan-related conflicts and the magnitude of displacement.

Consequently, this gap emphasizes the necessity for a more comprehensive and context-specific approach to elucidate the complex relationship between clan conflict and the scale of displacement, which constitutes the primary focus of this study.

## RESEARCH OBJECTIVES

The study examined four specific research objectives:

1. To identify specific regional variations in clan conflict-driven displacement in affected regions in Somalia.
2. To analyze the correlation between the frequency of clan conflicts and the scale of displacements across selected regions in Somalia.
3. To examine the extent to which clan conflict is a predictor of displacement.
4. To provide recommendations to mitigate clan conflict-driven displacement in Somalia.

### METHODS AND MATERIALS (RESEARCH METHODOLOGY)

This study utilized retrospective data. The Displacement Tracking Matrix (DTM) data in Somalia, produced by the International Organization for Migration (IOM), was employed; specifically, the study utilized data collected between April and October 2024 from 11 regions of Somalia (Bakool, Banadir, Bay, Galgaduud, Gedo, Hiraan, Lower Juba, Lower Shabelle, Middle Shabelle, Mudug and Nugaal).

The dataset was in Excel format and contained information on the study variables (clan conflict and displacement). The researchers conducted comprehensive data cleaning prior to data analysis, which involved checking for missing values, duplicates, completeness, relevance, and redundancy. After cleaning, the dataset contained 2,008 records of clan conflicts and 38,690 cases of displacement. There were no missing values, duplicates, or redundancies. Consequently, the dataset was deemed suitable for statistical analysis.

Data analysis was conducted utilizing statistical software packages for social science (SPSS) and an Excel spreadsheet to generate percentages, charts, tables, and statistical conclusions addressing the study objectives. The study employed descriptive and inferential statistics, including mean percentages, linear regression, and bivariate correlation analysis. The findings are presented in a systematic sequence of tables, numerical data, figures, and statistical inferences that address the study’s goals and objectives, as well as discussions, recommendations, and potential applications.

### ANALYSIS AND RESULTS OF THE STUDY

#### Regional variations in clan conflict-driven displacement in affected regions in Somalia

This research objective aims to provide the patterns and dynamics of clan conflict-driven displacement across different selected regions in Somalia. This analysis (Figure 1) will provide displacement across regions and how they vary regionally as well as specific insights for developing localized interventions and policy measures.

**Figure 1:**
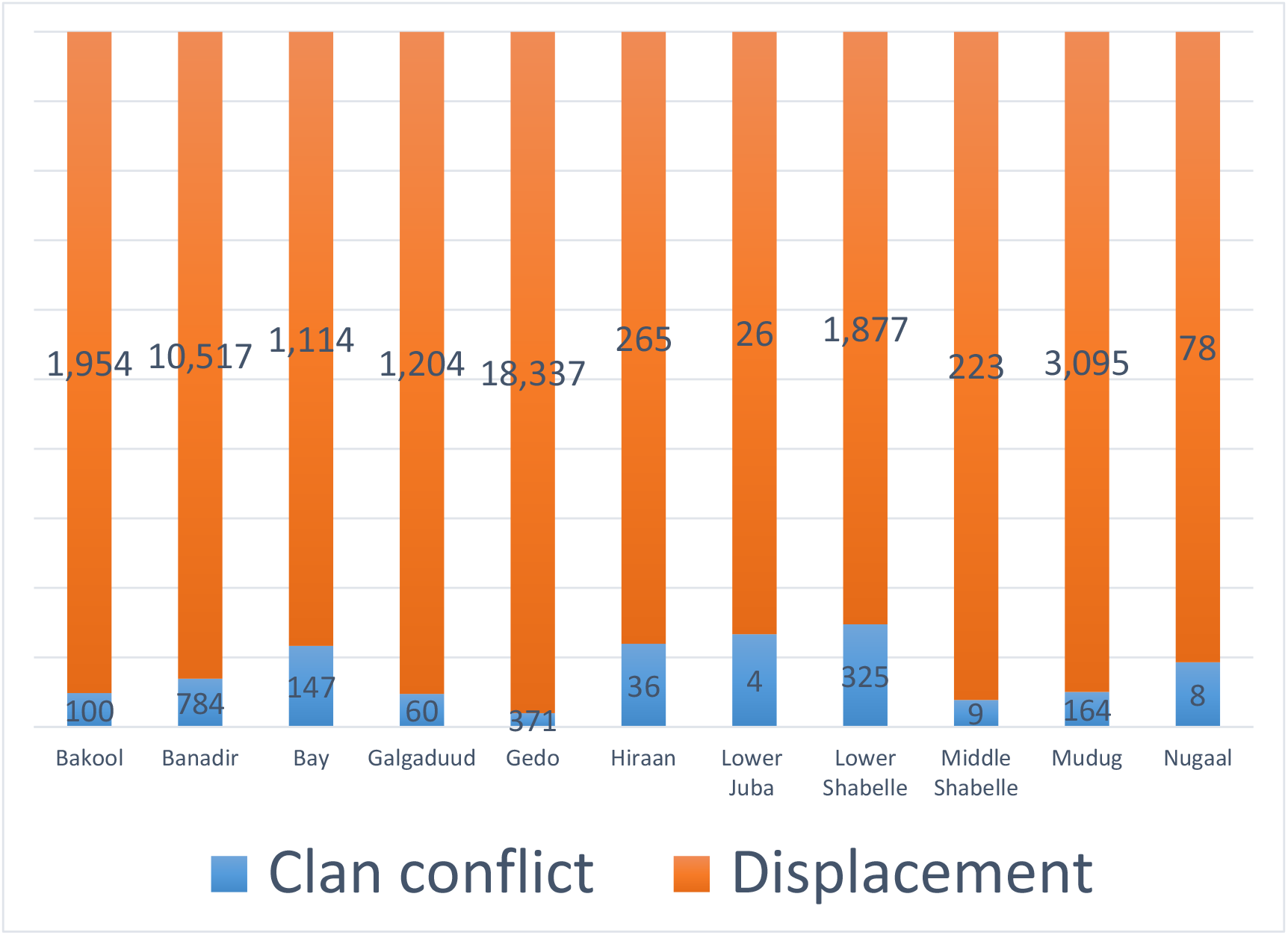
Regional variations in clan conflict-driven displacement.

The chart above illustrates the relationship between clan-based conflicts and population displacement across different regions in Somalia, showcasing notable variations. Gedo stands out as the most severely impacted area, experiencing an extraordinarily high level of clan-related strife at 18,387 incidents, resulting in the displacement of roughly 10,000 people. This striking difference underscores the strong link between escalated clan tensions and subsequent population displacement within this particular region.

Similarly, Banadir and Bay report significant clan-related conflicts, with 10,517 and 11,114 incidents respectively. However, their displacement figures are considerably lower than those in Gedo. This suggests that while clan conflicts are widespread, they don’t always result in proportionate displacement, possibly due to variations in local conditions or community resilience. In contrast, areas such as Hiraan, Lower Juba, and Middle Shabelle experience far fewer clan conflicts and displacements. Hiraan, for instance, records only 265 conflict incidents and minimal corresponding displacement. This indicates that these regions enjoy relative stability compared to more severely impacted areas.

In conclusion, the chart illustrates that while clan conflict frequently correlates with displacement, the magnitude and consequences vary considerably across regions. This nuanced comprehension is essential for tailoring interventions and support efforts in the most affected areas.

#### Correlation between the frequency of clan conflicts and the scale of displacements across selected regions in Somalia

This research objective analyzes the connection between clan conflict frequency and displacement magnitude in specific Somali regions. By examining this association, the research intends to measure how changes in clan conflict intensity affect displacement trends. Gaining insight into this relationship will show how clan conflicts influence displacement dynamics, enabling more focused interventions in areas impacted by conflict.

**Table 1:** Correlation between the frequency of clan conflicts and the scale of displacements.

| Correlations |  |  |  |
| --- | --- | --- | --- |
|  |  | Clan conflict | Displacement |
| Clan conflict | Pearson Correlation | 1 | .705* |
|  | Sig. (2-tailed) |  | .015 |
|  | N | 11 | 11 |
| Displacement | Pearson Correlation | .705* | 1 |
|  | Sig. (2-tailed) | .015 |  |
|  | N | 11 | 11 |

\*. Correlation is significant at the 0.05 level (2-tailed).

The researchers used Correlation Coefficient analysis to assess the relationship between the frequency of clan conflicts and the scale of displacements. The table above presents the results of a correlation analysis. The Correlation Coefficient of 0.705 indicates a strong positive correlation between clan conflict and displacement, suggesting that as one variable increases, the other tends to increase as well. Furthermore, the P-value is less than the conventional alpha level of 0.05, indicating that the correlation is statistically significant. This finding allows for the confident assertion that the observed relationship is unlikely to be attributable to random chance.

In conclusion, there is a significant positive correlation between clan conflict and displacement, indicating that higher levels of clan conflict are associated with increased levels of displacement.

#### Examining the extent to which clan conflict is a predictor of displacement

The above research objective aims to assess how well clan-based conflicts can predict population displacement in Somalia, utilizing Linear Regression Analysis. The research aims to assess and quantify the impact and relevance of clan conflict as a predictor factor in displacement by analyzing the relationship between these two variables under the below tables.

**Table 2:** Model summary of linear regression analysis.

| Model Summary |  |  |  |  |
| --- | --- | --- | --- | --- |
| Model | R | R Square | Adjusted R Square | Std. Error of the Estimate |
| 1 | .705<sup>a</sup> | .497 | .441 | 4298.106 |

a. Predictors: (Constant), Clan conflict

The model summary provides insights into the relationship between clan conflict and displacement. The correlation coefficient (R) is 0.705, indicating a strong positive relationship; as clan conflict increases, displacement also tends to increase. The R Square value is 0.497, signifying that approximately 49.7% of the variance in displacement can be explained by clan conflict. This indicates that while clan conflict is a significant predictor of displacement, other factors likely contribute to the variations in displacement that are not captured by this model. In summary, the data indicates a significant relationship between clan conflict and displacement.

**Table 3:** ANOVA of linear regression analysis.

| ANOVA<sup>a</sup> |  |  |  |  |  |  |
| --- | --- | --- | --- | --- | --- | --- |
| Model |  | Sum of Squares | df | Mean Square | F | Sig. |
| 1 | Regression | 164243768.1 | 1 | 164243768.1 | 8.891 | .015<sup>b</sup> |
|  | Residual | 166263404.1 | 9 | 18473711.57 |  |  |
|  | Total | 330507172.2 | 10 |  |  |  |

a. Dependent Variable: Displacement
b. Predictors: (Constant), Clan conflict

The ANOVA table above provides insights into the overall significance of the regression model used to predict displacement based on clan conflict. The ANOVA results indicate that the regression model significantly explains the variance in displacement related to clan conflict. The F-statistic of 8.891 and the p-value of 0.015 suggest that the model is statistically significant, reinforcing the conclusion that clan conflict has a meaningful effect on displacement.

**Table 4:** Coefficients of linear regression analysis.

| Coefficients<sup>a</sup> |  |  |  |  |  |  |
| --- | --- | --- | --- | --- | --- | --- |
| Model |  | Unstandardized Coefficients |  | Standardized Coefficients | t | Sig. |
|  |  | B | Std. Error | Beta |  |  |
| 1 | (Constant) | 376.798 | 1669.954 |  | .226 | .827 |
|  | Clan conflict | 17.204 | 5.770 | .705 | 2.982 | .015 |

a. Dependent Variable: Displacement

The coefficients table above provides detailed information about the relationship between clan conflict and displacement, with “Displacement” as the dependent variable. The Standardized Coefficients (Beta) value for clan conflict is 0.705, indicating that clan conflict has a strong effect on the scale of displacement. Moreover, the significance value for clan conflict is 0.015. This p-value indicates that the relationship between clan conflict and displacement is statistically significant (typically, a p-value less than 0.05 is considered significant). This suggests that clan conflict is a meaningful predictor of displacement.

In summary, the coefficients table indicates a statistically significant and positive relationship between clan conflict and displacement. For each unit increase in clan conflict, displacement is expected to increase by about 17.204 units (unstandardized coefficient). The results suggest that addressing clan conflict could be crucial in managing and mitigating displacement issues.

## DISCUSSION

This study provides compelling evidence for the significant association between clan-based conflicts and population displacement in Somalia. The study’s findings, encompassing geographic variations, robust correlations, and predictive models, collectively elucidate the critical influence of conflict dynamics on displacement patterns. To mitigate the humanitarian and societal ramifications of displacement, it is imperative to address the root causes of these conflicts through targeted, conflict-sensitive, and governance-oriented interventions. By leveraging these findings, policymakers and stakeholders can develop more effective strategies to address the complex issues surrounding clan conflict and displacement in Somalia. This is in line with the research elucidates substantial regional disparities in displacement across selected regions in Somalia.

The Gedo region was identified as the most severely affected, experiencing 18,387 clan-related conflicts that resulted in approximately 10,000 displacements. These regional variations underscore the need for tailored interventions addressing area-specific issues while enhancing resilience in the most impacted communities. The study findings on the scale of displacement demonstrates a substantial figure due to clan conflict, amounting to 38,690 over seven months. This aligns with data from the Somali humanitarian needs and response plan, which indicates that over 6 million Somalis are affected by the crisis and require urgent humanitarian assistance to prevent loss of life (UNOCHA, 2024), The magnitude of displacement observed in this research aligns with another study, “Environment-induced internal displacement: The case of Somalia,” which indicates that internally displaced persons (IDPs) constitute 56% of all forced migration. Significantly, this displacement persists for an average of 20 years for refugees and over 10 years for most IDPs. Moreover, the UNHCR (2020) global trends report on forced displacement revealed that nearly 80% of refugees and IDPs reside in developing nations. Somalia, consistently ranking among the top 10 countries of origin for cross-border displacement over the past decade, placed fifth globally in terms of forcibly displaced populations, with approximately 2.6 million IDPs (Momeni et al., 2022).

The robust positive correlation between clan conflicts and displacement (r = 0.705, p = 0.015) suggests that regions experiencing higher frequencies of conflicts tend to exhibit elevated levels of population displacement. This research somehow aligns with another study examining “scarce resources and inter-clan conflict in selected districts in Madera County, Northeastern Kenya.” The findings from that study revealed a strong correlation between resource scarcity and inter-clan conflicts. Specifically, the linear correlation coefficient analysis demonstrated a significant relationship between land scarcity and inter-clan conflict (r=0.803, Sig 0.000). Similarly, water scarcity exhibited a strong connection to all aspects of inter-clan conflict (r=O.860, SlG=O.000). Comparable results were observed for pasture scarcity (r=0.774, SIG =0.000), indicating its substantial impact on inter-clan tensions(Shale, 2011).

The predictive strength of clan conflict on displacement is further substantiated by linear regression analysis. The model demonstrates that clan conflict intensity accounts for approximately half (49.7%) of the variability in displacement, as indicated by an R^2^ value of 0.497. The regression model’s statistical significance (F = 8.891, p = 0.015) underscores the critical importance of addressing clan conflict as part of comprehensive strategies aimed at mitigating displacement. The findings align with the research titled “Scarce Resources and Inter-clan Conflict in Selected Districts in Madera County, Northeastern Kenya.” The results of regression analysis indicated that the model’s independent variables exhibited a statistically significant impact on variations in the dependent variable (inter-clan conflict) (F=1019.69, sig. =0.000). This led researchers to conclude that resource scarcity constituted a significant factor in elucidating the high incidence of inter-clan conflicts within Madera County (Shale, 2011).

### RECOMMENDATIONS

Given the findings and analysis demonstrated above, the study suggests the following recommendations:

1. To mitigate the magnitude of displacement resulting from clan conflicts, the researchers strongly advocate for the implementation of community-based resilience strategies, particularly in regions characterized by high conflict and displacement levels. This can be achieved through the enhancement of social cohesion, local dispute resolution mechanisms, and economic opportunities. Such measures will contribute to reducing the community’s vulnerability to displacement.
2. The study recommends government and humanitarian actors to establish and implement early warning systems to monitor and predict clan conflicts in high-risk regions, thereby facilitating timely interventions to mitigate large-scale displacement.
3. The researchers suggests that data-driven policy making can be enhanced through the utilization of statistical findings, such as the identified strong positive correlation between clan conflicts and displacement. These insights can be applied to inform policy decisions and develop interventions aimed at mitigating the effects of clan conflicts in Somalia.
4. The research recommends that the implementation of programs offering sustainable income-generating opportunities for displaced populations is advisable, particularly in regions such as Gedo where displacement significantly impacts economic stability. These initiatives may encompass skills development and access to microfinance services, with the objective of supporting viable livelihoods.
5. The research further suggests the necessity of fostering collaborations among governmental entities, community organizations, international institutions, and non-governmental organizations to establish a coordinated approach for addressing clan-based conflicts and population displacement. This strategy aims to optimize the utilization of available resources and expertise.
6. Implement conflict-aware strategies by focusing on areas such as Gedo, Banadir, and Bay, where clan disputes and population displacement are most prevalent. Provide customized assistance that addresses the unique challenges and susceptibilities faced by these communities.

## CONCLUSION

The study into clan conflict dynamics and their role in driving displacement has yielded significant insights regarding the relationship between clan conflict and the magnitude of displacement, as well as elucidated the regional variation of displacement.

The findings indicate that Gedo experiences the highest incidence of clan conflicts and resulting population displacements, making it the most severely affected region. In contrast, areas like Hiraan and Middle Shabelle are less impacted by warfare and have witnessed minimal population movement. The correlation analysis has verified that increased clan disputes significantly contribute to population displacement, as evidenced by a statistically significant positive relationship between the number of clan conflicts and the magnitude of displacement (r = 0.705, p < 0.05). The research underscores the importance of implementing region-specific interventions and strategies to mitigate the impact of these conflicts.

Finally, addressing this issue requires a comprehensive approach that incorporates conflict prevention, community awareness, enhancement of community resilience, and provision of livelihood support, particularly in the most affected regions. To develop and implement sustainable solutions that promote stability and mitigate displacement resulting from clan conflicts in Somalia, policymakers and stakeholders must utilize evidence-based insights.

## Data availability statement

The data underpinning the findings of this study are sourced from third-party data provided by the International Organization for Migration (IOM). The minimal dataset utilized to derive the conclusions presented in this manuscript is obtained from the Displacement Tracking Matrix (DTM) data in Somalia, covering the period from April to October 2024. These data are publicly accessible at Displacement.iom.int: https://displacement.iom.int/somalia/ or Global DTM: https://dtm.iom.int/somalia.

## REFERENCES

Abdikarim Mohaidin, A. (2011). Civil war and poverty enhancement of selected internal displaced people (idp) camps in Mogadishu Somalia.

Abdirahman, M. I. (2016). Ethnic conflict and socio-economic development of selected house holders in Mismayo District of Somalia.

Adan, M. A., & Orodho, J. A. (2013). Effects of Inter-Clan Conflicts On Quality School Outcomes in Secondary Schools among Nomadic Communities in Mandera County, Kenya. IOSR Journal of Research & Method in Education (IOSR-JRME) e-ISSN, 2320-7388.

Addow, M. H. (2024). The lives and livelihoods of forcibly displaced people in Mogadishu, Somalia.

Ahmedweli Mohamud, M. (2012). Conflict and peace process in Somalia Kampala International University, College of Humanities and Social Sciences].

Alio, H. M. (2012). Inter-clan conflict in Mandera District: a case of the Garre and Murulle, 2004-2009 University of Nairobi, Kenya].

Aymerich, O., & Zeyneloglu, S. (2023). Sense of Community and Conflict-induced Displacement: A Study among Iraqi IDPs in Camps. Journal of Internal Displacement, 13(1), 20–36.

Barrow, I. H. (2020). Inter-clan conflicts in Somalia: When peace happen (case study Baidoa District, Bay Region). International Journal of Human Resource Studies, 10(4), 111–111.

Blair, C. W. (2024). Dynamics of Internal Displacement and Conflict in Somalia. Available at SSRN.

Drumtra, J. (2014). Internal displacement in Somalia. Brookings Institution Washington, DC.

Farah, I., Hussein, A., & Lind, J. (2002). Deegaan, politics and war in Somalia. Scarcity and surfeit: The ecology of Africa’s conflicts, 321–356.

Fellin, M. (2013). The historical impact of western colonial and imperial policies and interventions on conflict and internal displacement in Somalia. Journal of Internal Displacement, 3(2), 41–62.

Gundel, J. (2009). Clans in Somalia: Report on a lecture by Joakim Gundel. COI Workshop Vienna,

Happi, R. A. (2016). Influence Of Somali Clans’ Conflict On Teachers’ Retention In Public Primary Schools In Banisa Subcounty, Mandera County, Kenya University Of Nairobi].

Hussein, M. (2022). Inter-Clan Conflicts In the Somali Region of Ethiopia: A Case Study Of the Conflicts between the Dagodia and Baydisle Clans

Jama, A. S., Samanter, M., Muturi, W., & Mberia, H. (2018). Causes of land conflict in Puntland state of Somalia the case for Garowe city. International Journal of Advanced Research and Development, 3(3), 07–16.

Jibril, S. M. (2013). Intra-Clan Conflict between Ida’gale and Habaryonis and the Role of Traditional Leaders in Pastoral Conflict Resolution in Aware District, Somali Region of Ethiopia. Asian Journal of Humanities and Social Studies, 1(3).

Kamta, F. N., & Scheffran, J. (2022). A social network analysis of internally displaced communities in northeast Nigeria: potential conflicts with host communities in the Lake Chad region. GeoJournal, 87(5), 4251–4268.

Kinyoki, D. K., Moloney, G. M., Uthman, O. A., Kandala, N.-B., Odundo, E. O., Noor, A. M., & Berkley, J. A. (2017). Conflict in Somalia: impact on child undernutrition. BMJ global health, 2(2), e000262.

Le Sage, A., & Majid, N. (2002). The livelihoods gap: Responding to the economic dynamics of vulnerability in Somalia. Disasters, 26(1), 10–27.

Mohamed, A. (2018). Governance, clan conflicts and socio-economic development in Baidoa District, Somalia Kampala International University, College of Humanities and Social Sciences].

Mohamed, A. A. (2018). Assessment of conflict dynamics in Somali national regional state of Ethiopia. Journal of public policy and administration, 2(4), 40–48.

Mohamed, M. S., & Samantar, M. Factors Affecting To Local Integration For Internally Displaced Persons In Garowe, Somalia.

Momeni, R., Bircan, T., & King, R. (2022). Environment-induced internal displacement. The case.

Muna Mohamed, Y. (2020). Internal displacement and women rights among the residents of Deeq Rabbi Camp, Mogadishu, Somalia Kampala International University: College of Humanities and Social Sciences].

Mustafe Mahamoud, A. (2017). Disasters and forced displacement in Rajo IDPS Mogadishu Somalia Kampala International University, College of Humanities and Social Sciences].

Ongaga, J. N. (2017). YOUNG INITIATES INVOLVEMENT IN INTER-CLAN CONFLICT MANAGEMENT AND RESOLUTION IN TODONYANG SUB-LOCATION, TURKANA COUNTY, 2013 TO 2017 Kenyatta University].

Osman, A. A., & Abebe, G. K. (2023). Rural displacement and its implications on livelihoods and food insecurity: the case of inter-riverine communities in Somalia. Agriculture, 13(7), 1444.

Robinson, C., Zimmerman, L., & Checchi, F. (2014). Internal and external displacement among populations of Southern and Central Somalia Affected by Severe Food Insecurity and Famine during 2010–2012. DC: FEWS NET.

Sahal, M. A. (2016). Factors Influencing Perennial Sub-clan Conflict in Garissa County, Kenya University of Nairobi].

Salad, B. H. (2015). Factors influencing inter-clan conflict in northern Kenya: a case of Wajir county University of Nairobi].

Sate, G. R., & Konso Zone, S. N. Conflict and displacement in.

Schon, J. (2015). Focus on the forest, not the trees: A changepoint model of forced displacement. Journal of refugee studies, 28(4), 437–467.

Shale, I. A. (2011). Scarce resources and inter-clan conflict in selected districts in Mandera county, north eastern Kenya Kampala International University, College of Humanities and Social Sciences].

UNOCHA. (2024). SOMALIA - HUMANITARIAN NEEDS AND RESPONSE PLAN.

Yigzaw, G. S., & Abitew, E. B. (2019). Causes and impacts of internal displacement in Ethiopia. African Journal of Social Work, 9(2), 32–41.

